# A comparative analysis of COVID-19 mortality rate across the globe: An extensive analysis of the associated factors

**DOI:** 10.1101/2020.12.22.20248696

**Authors:** Vineet Jain, Nusrat Nabi, Kailash Chandra, Sana Irshad, Varun kashyap, Sunil Kohli, Arun Gupta

## Abstract

**Background:** The vast variation in COVID 19 mortality across the globe draws attention to potential risk factors other than the patient characteristics that determine COVID-19 mortality.

**Subjects and Methods:** We have quantified and analyzed one of the broadest set of clinical factors associated with COVID-19-related death, ranging from disease related co-morbities, socioeconomic factors, healthcare capacity and government policy and interventions. Data for population, total cases, total COVID mortality, tests done, and GDP per capita were extracted from the worldometers database. Datasets for health expenditure by government, hospital beds, rural population, prevalence of smoking, prevalence of overweight population, deaths due to communicable disease and incidence of malaria were extracted from the World Bank website. Prevalence of diabetes was retrieved from the indexmundi rankings. The average population age, 60+ population, delay in lockdown, population density and BCG data were also included for analysis. The COVID-19 mortality per million and its associated factors were retrieved for 56 countries across the globe. Quantitative analysis was done at the global as well as continent level. All the countries included in the study were categorized continent and region wise for comparative analysis determining the correlation between COVID 19 mortality and the aforementioned factors.

**Results:** There was significant association found between mortality per million and 60+ population of country, average age, prevalence of diabetes mellitus, and case fatality rate with correlation and p value (p) of 0.422 (p 0.009), 0.386 (p 0.0186), −0.384 (p 0.019) and 0.753 (p 0.000) respectively at 95% CI.

**Conclusion:** The study observations will serve as a evidence based management strategy for generating predictive model for COVID-19 infection and mortality rate.

## Background

WHO declared COVID-19 as a pandemic on March 11, 2020, due to worldwide alarming levels of spread and severity of COVID-19 (Bedford *et al*., 2020). One of the questions studied extensively in the pandemic is the huge variation in the COVID-19 mortality rate across the globe, from over 16% in France and Belgium to less than 0.1% in Singapore and Qatar (Liang *et al*., 2020). Such wide variation implies that there are factors other than patient characteristics that determine COVID-19 mortality, such as population dynamics, social and economical structure and government response.

Studies have well established that COVID-19 mortality is positively associated with the patient characteristics such as the increasing age, obesity and underlying comorbidities, like hypertension, diabetes, and coronary heart disease (Chen *et al*., 2020; Guan *et al*., 2020; Zhou *et al*., 2020). Studies have also suggested that increasing COVID-19 testing has been associated with a slow viral spread (Peto, 2020). Some epidemiologists have attributed the government response to containing the spread of the virus (Iacobucci, 2020). Countries vary widely in terms of demographic profile, economic status, health infrastructure and ability to respond promptly to the disease outbreaks. Hence, it is crucial to design and implement rational strategies for the safety of patients, healthcare professionals and general public and to ensure all nations work together cohesively to emerge out successfully from the pandemic. We have done a diverse analysis to apply the evidence based explanation for the impact of socioeconomic factors, healthcare capacity, government policy and interventions, and the disease related risk factors on COVID-19 associated mortality. In the current study, we have used the COVID mortality per million population, a stable parameter as the yardstick to correlate the COVID 19 mortality with the associated factors, and not the usual case fatality rate (CFR is the number of deaths per 100 confirmed cases) because the CFR depends on the testing capacity of a country and hence does not reflect the true mortality.

### Subjects and Method

This is a worldwide cross-sectional study. The selection of the risk-related factors of COVID-19 was based on review of the literature and essentially based on data availability and completeness. The data for the study was extracted as on August 21, 2020. Data for total population, total cases, total COVID deaths, tests done, and GDP per capita (nominal in USD) were extracted from the Worldometers Coronavirus Statistics website (*Coronavirus Update (Live) - Worldometer*, no date). Datasets for health expenditure as percentage of GDP by government, number of hospital bed numbers (per 1000 people), percentage of rural population, prevalence of smoking (15+ age), percentage of overweight adult population, percentage of deaths due to communicable disease & other nutrition conditions and incidence of malaria (per 1000 population) were extracted from the World Bank Coronavirus website (*World Bank Open Data* | *Data*, no date). Prevalence of diabetes mellitus (percentage of adult population) was retrieved from the indexmundi rankings (Index mundi, no date). The key findings from world population prospects 2017 were used for percentage of 60+ population and the average population age (United Nations, 2017). Delay in lockdown (with regards to the first case in the country)(Wikipedia, 2020), population density per square meter(*World Population Growth - Our World in Data*, no date) and current BCG vaccination policy(*BCG World Atlas, 3rd edition*, no date) were also included for analysis. Out of about 185 affected countries, only 56 countries with total confirmed cases more than 25000 (as on August 21, 2020) were selected for analysis in this study. COVID-19 mortality and associated factors were studied at the global level and between the various continents and regions; continent 1; Asia, continent 2; Europe, continent 3; North America, continent 4; South America, continent 5; Africa and region 6; Middle East.

### Statistical analysis

The risk-related factors with different units of measurement were standardized. The mortality per million population was calculated and compared with each parameter included in the study for any statistically significant association. The factors were examined using the Pearson correlation matrix and Percentage while comparison of continent we use one way analysis of variance method (ANOVA).

## Results

In this study, we observed few parameters significantly associated with COVID mortality across the globe. The results observed at the global level are summarized in **Table 1**. There was significant association found between mortality per million and 60+ population of country, average age, prevalence of diabetes mellitus, and case fatality rate with the correlation and p value (p) of 0.422 (p 0.009), 0.386 (p 0.0186), −0.384 (p 0.019),and 0.753 (p 0.000) respectively at 95% CI. The Pearson correlation analysis in each continent is revealed in **Tables 2-6**. The results revealed some significant correlations in the continent and region wise analysis, however, few countries in the same region also behaved differently which is detailed in the discussion.

**Table 1:** Correlation of COVID-19 mortality per million with the possible predictors, globally.

| Predictors | Number of X (Mortality per mil) Y (predictors) Pairs | Pearson r | P value (two-tailed) |
| --- | --- | --- | --- |
| Test per million | 56 | -0.014 | 0.934 |
| GDP per capita (nominal) | 56 | 0.324 | 0.051 |
| Health expenditure per GDP | 56 | -0.152 | 0.371 |
| 60+ population | 56 | 0.422 | **0.009** |
| Rural population % | 56 | -0.194 | 0.251 |
| Average age | 56 | 0.386 | **0.018** |
| PDM (%) | 56 | -0.384 | **0.019** |
| Population (million) | 56 | -0.124 | 0.464 |
| Population density (people in per Sqmt) | 56 | -0.165 | 0.330 |
| Total test (n) | 56 | 0.051 | 0.767 |
| Total cases (n) | 56 | 0.261 | 0.118 |
| Cases per million | 56 | 0.041 | 0.812 |
| Case fatality rate (%) | 56 | 0.753 | **0.000** |
| BCG vaccination | 56 | -0.088 | 0.604 |
| Overweight (% of adult) | 56 | 0.183 | 0.278 |
| Smoking prevalence | 56 | 0.137 | 0.425 |
| Beds per 1000 people | 56 | 0.101 | 0.554 |
| Nutrition condition (% of total) | 56 | -0.210 | 0.213 |

**Dependent Variable:** mortality per mil, PDM: Prevalence of diabetes mellitus.

**Table 2:** Correlation of COVID-19 mortality per million with the possible predictors in Asian continent.

| Predictors | Number of X (Mortality per mil) Y (predictors) Pairs | Pearson r | P value (two-tailed) |
| --- | --- | --- | --- |
| Test per million | 22 | -0.071 | 0.755 |
| GDP per capita (nominal) | 22 | -0.228 | 0.307 |
| Health expenditure per GDP | 22 | -0.197 | 0.379 |
| 60+ population | 22 | -0.100 | 0.657 |
| Rural population % | 22 | -0.110 | 0.635 |
| Average age | 22 | -0.011 | 0.962 |
| PDM (%) | 22 | -0.107 | 0.634 |
| Population (million) | 22 | -0.322 | 0.143 |
| Population density (people per Sqmt) | 22 | -0.245 | 0.272 |
| Total test (n) | 22 | -0.270 | 0.224 |
| Total cases (n) | 22 | -0.129 | 0.566 |
| Cases per million | 22 | 0.253 | 0.256 |
| Case fatality rate (%) | 22 | 0.166 | 0.461 |
| BCG vaccination | 22 | 0.132 | 0.700 |
| Overweight (% of adult) | 22 | **0.500** | **0.018** |
| Smoking prevalence | 22 | -0.132 | 0.578 |
| Beds per 1000 people | 22 | -0.007 | 0.974 |
| Cause of death due to communicable disease and others nutrition conditions (% of total) | 22 | **-0.436** | **0.043** |

Dependent Variable: mortality per mil

**Table 3:** Correlation of COVID-19 mortality per million with the possible predictors in European continent.

| Predictors | Number of X (Mortality per mil) Y<br>(predictors) Pairs | Pearson r | P value (two-tailed) |
| --- | --- | --- | --- |
| Test per million | 16 | 0.406 | 0.118 |
| GDP per capita (nominal) | 16 | -0.228 | 0.307 |
| Health expenditure per GDP | 16 | -0.197 | 0.379 |
| 60+ population | 16 | -0.100 | 0.657 |
| Rural population % | 16 | -0.110 | 0.635 |
| Average age | 16 | -0.011 | 0.962 |
| PDM (%) | 16 | -0.107 | 0.634 |
| Population (million) | 16 | -0.168 | 0.534 |
| Population density (people per Sqmt) | 16 | 0.509 | 0.044 |
| Total test (n) | 16 | -0.129 | 0.566 |
| Total cases (n) | 16 | -0.129 | 0.566 |
| Cases per million | 16 | 0.253 | 0.256 |
| Case fatality rate (%) | 16 | 0.166 | 0.461 |
| BCG vaccination | 16 | 0.132 | 0.700 |
| Overweight (% of adult) | 16 | -0.397 | 0.256 |
| Smoking prevalence | 16 | -0.339 | 0.198 |
| Beds per 1000 people | 16 | -0.503 | 0.056 |
| Cause of death due to communicable disease and others nutrition conditions (% of total) | 16 | 0.455 | 0.077 |

Dependent Variable: mortality per mil

**Table 4:** Correlation of COVID-19 mortality per million with the possible predictors in North America.

| Predictors | Number of X (Mortality per mil) Y<br>(predictors) Pairs | Pearson r | P value<br>(two-tailed) |
| --- | --- | --- | --- |
| Test per million | 07 | 0.638 | 0.123 |
| GDP per capita (nominal) | 07 | 0.481 | 0.059 |
| Health expenditure per GDP | 07 | **0.539** | **0.031** |
| 60+ population | 07 | 0.362 | 0.168 |
| Rural population % | 07 | **-0.665** | **0.005** |
| Average age | 07 | **0.657** | **0.006** |
| PDM (%) | 07 | -0.533 | 0.034 |
| Population (million) | 07 | 0.772 | 0.042 |
| Population density (people per Sqmt) | 07 | -0.701 | 0.080 |
| Total test (n) | 07 | 0.659 | 0.107 |
| Total cases (n) | 07 | 0.002 | 0.993 |
| Cases per million | 07 | 0.479 | 0.060 |
| Case fatality rate (%) | 07 | 0.870 | 0.000 |
| BCG vaccination | 07 | -0.479 | 0.277 |
| Overweight (% of adult) | 07 | **-0.031** | **0.909** |
| Smoking prevalence | 07 | -0.339 | 0.198 |
| Beds per 1000 people | 07 | -0.503 | 0.056 |
| Cause of death due to communicable disease and others nutrition conditions (% of total) | 07 | **0.455** | **0.077** |

Dependent Variable: mortality per mil

**Table 5:** Correlation of COVID-19 mortality per million with the possible predictors in South America.

| Predictors | Number of X (Mortality per mil) Y<br>(predictors) Pairs | Pearson r | P value (two-tailed) |
| --- | --- | --- | --- |
| Test per million | 07 | **0.789** | **0.035** |
| GDP per capita (nominal) | 07 | 0.627 | 0.132 |
| Health expenditure per GDP | 07 | 0.593 | 0.161 |
| 60+ population | 07 | 0.531 | 0.220 |
| Rural population % | 07 | -0.506 | 0.246 |
| Average age | 07 | 0.732 | 0.061 |
| PDM (%) | 07 | 0.487 | 0.268 |
| Population (million) | 07 | 0.144 | 0.758 |
| Population density (people per Sqmt) | 07 | 0.001 | 0.758 |
| Total test (n) | 07 | 0.336 | 0.462 |
| Total cases (n) | 07 | 0.691 | 0.086 |
| Cases per million | 07 | 0.606 | 0.149 |
| Case fatality rate (%) | 07 | 0.296 | 0.519 |
| Overweight (% of adult) | 07 | 0.733 | 0.061 |
| Smoking prevalence | 07 | 0.528 | 0.281 |
| Beds per 1000 people | 07 | 0.714 | 0.071 |
| Cause of death due to communicable disease and others nutrition conditions (% of total) | 07 | -0.646 | 0.117 |

Dependent Variable: mortality per mil

**Table 6:** Correlation of COVID-19 mortality per million with the possible predictors in Africa.

| Predictors | Number of X (Mortality per mil) Y (predictors) Pairs | Pearson r | P value (two-tailed) |
| --- | --- | --- | --- |
| Test per million | 04 | 0.900 | 0.100 |
| GDP per capita (nominal) | 04 | -0.053 | 0.910 |
| Health expenditure per GDP | 04 | -0.322 | 0.481 |
| 60+ population | 04 | -0.122 | 0.795 |
| Rural population % | 04 | 0.082 | 0.861 |
| Average age | 04 | -0.050 | 0.916 |
| PDM (%) | 04 | 0.416 | 0.353 |
| Population (million) | 04 | -0.371 | 0.629 |
| Population density (people per Sqmt) | 04 | -0.792 | 0.208 |
| Total test (n) | 04 | 0.929 | 0.071 |
| Total cases (n) | 04 | 0.282 | 0.539 |
| Cases per million | 04 | **0.825** | **0.022** |
| Case fatality rate (%) | 04 | 0.371 | 0.412 |
| BCG vaccination | 04 | -0.494 | 0.397 |
| Overweight (% of adult) | 04 | -0.216 | 0.642 |
| Smoking prevalence | 04 | -0.008 | 0.989 |
| Beds per 1000 people | 04 | -0.512 | 0.240 |
| Cause of death due to communicable disease and others nutrition conditions (% of total) | 04 | 0.012 | 0.980 |

Dependent Variable: mortality per mil

**Table 7:** Correlation of COVID-19 mortality per million with the possible predictors in Middle East countries.

| Predictors | Number of X (Mortality per mil) Y (predictors) Pairs | Pearson r | P value (two-tailed) |
| --- | --- | --- | --- |
| Test per million | 11 | -0.398 | 0.225 |
| GDP per capita (nominal) | 11 | -0.437 | 0.178 |
| Health expenditure per GDP | 11 | 0.616 | 0.043 |
| 60+ population | 11 | 0.081 | 0.814 |
| Rural population % | 11 | -0.014 | 0.814 |
| Average age | 11 | -0.313 | 0.348 |
| PDM (%) | 11 | 0.647 | 0.031 |
| Population (million) | 11 | 0.175 | 0.607 |
| Population density (people per Sqmt) | 11 | -0.053 | 0.678 |
| Total test (n) | 11 | -0.192 | 0.572 |
| Total cases (n) | 11 | 0.568 | 0.070 |
| Cases per million | 11 | **-0.144** | **0.673** |
| Case fatality rate (%) | 11 | 0.451 | 0.164 |
| Overweight (% of adult) | 11 | -0.380 | 0.279 |
| Smoking prevalence | 11 | -0.732 | 0.016 |
| Beds per 1000 people | 11 | -0.140 | 0.682 |
| Cause of death due to communicable disease and others nutrition conditions (% of total) | 11 | 0.350 | 0.292 |

Dependent Variable: mortality per mil

## Discussion

Recording mortality itself depends on the COVID status of a patient (positive or suspected), method used for testing (standard RTPCR or antigen based), method of calculating mortality (case fatality rate or mortality per million population) and the places from where the patients are picked (hospitals, care homes or labs). Extensive and systematic analysis of interactions between various factors associated with COVID outcomes and mortality form the cornerstone of COVID 19 management. We have studied one of the broadest ranges of factors with COVID-19 mortality for any possible correlation which can explain the COVID 19 disease pattern.

### Age

Elderly people are more susceptible to the risk of getting infected with this disease, resulting in increased mortality(Verity *et al*., 2020). With increasing age the deficiency of T-cell and B-cell function rises, and there is overproduction of type 2 cytokines(Zhou *et al*., 2020). This usually adds up to the viral replication and stretch the period of pro-inflammatory responses which result in poor health conditions (Zhou *et al*., 2020). Verity et al. (2020) observed that the case fatality rate for those under age 60 was 1.4%, however, it increased drastically to 4.5% for people aged over 60 years old and to 13.4% for 80 years and above (Verity *et al*., 2020). Moreover, the average age of people dying of COVID-19 was found to be 68 years, while, the average age for recovered patients was 51 years(Chen *et al*., 2020).

In the present study we observed a positive as well as statistically significant global association of 60+ population percentage with COVID mortality (r=0.422; p=0.009). Further comparing the data between the various continents, it was found that higher ‘60+ population’, such as in Europe (25-29%) was positively associated with COVID 19 morality. Whereas, regions with younger population like the Asia, Africa and Middle-East witnessed fewer deaths. Exceptions to this association were South America and North America (except USA, Canada) which had only 10-15% ‘60+ population’ but experienced higher mortality. On the other hand, individual countries from Asia, like Japan (33%) and Singapore (20%) with high ‘60+ population’ had less mortality.

### Obesity

Increase in obesity is associated with higher risk of COVID-19 mortality and is implicated in severe outcomes comprising of invasive mechanical ventilation, acute pneumonia, increased hospitalizations and COVID 19 mortality(Liang *et al*., 2020). Some of the proposed etiopathogenesis include restrictive lung disease, lipotoxicity and induction of a pro-inflammatory state associated with the increased BMI (Sattar, McInnes and McMurray, 2020). Similar results with significantly increased risk of mortality were documented in a study conducted in the overweight and obese groups compared with the normal BMI group, after controlling for age, gender, diabetes, hypertension, and qSOFA score (Nakeshbandi *et al*., 2020).

The findings of our study revealed a positive but statistically non-significant association of percentage of overweight adults with mortality at the global level. However, significantly less COVID 19 mortality was observed in Asia (r=0.5; p= 0.018) which has less percentage of obese adults. Whereas, South America with high prevalence of overweight; reported a corresponding high mortality (r=0.733; p=0.061). Countries like Nigeria (29%) and Ghana (32%) with less obesity had less mortality; and South Africa with 54% obesity had high mortality. However, Middle East countries like Egypt with an obesity of 64% had unusually low mortality rate.

### Population Density

Areas which are densely populated usher direct contacts among the residents, forming embryonic hotspots resulting in pandemic escalation. Looking at it the other way, such heavily populated areas may be a step ahead in providing quality and accessible health care facilities and administering better social distancing policies (You, Wu and Guo, 2020). A US based study revealed that areas with large population have greater COVID-19 infection and fatality rates. However, the observation was reversed after controlling for population size and other confounding variables, resulting in significant negative association of infection and mortality rates with the population density(Hamidi, Ewing and Sabouri, 2020).

The study of population density across the globe reflected a negative (non-significant) association with COVID mortality. We observed a vast difference in mortality when comparing countries with similar population density such as India (394) and Belgium (379) with a contrasting mortality rate of 40 and 906 respectively. Similar contrasting mortality pattern has been documented for other countries with matching population density, like UK and Pakistan and; Italy and Qatar. Our data was unable to find a predictive pattern for population density variable. On one hand we had densely populated Asia with less mortality and sparsely populated American continent on the other hand with a high mortality. However, Europe with some dense populated countries like Netherland, Belgium, UK and Italy; did show a positive association with mortality rate (r=0.509; p=0.044).

### Rural Population

Lack of health infra-structure, basic health services, testing facilities and accessibility to health care as well as low socio-economic status are important factors positively associated with the mortality rate in the rural areas. However, the less dense population setup and low human and goods exchange facilitates the social distancing, hence the spread of the disease. However, generalizing this rule may again hamper the monitoring of COVID-19 spread, as on one hand many rural communities have become virus hotspots and on the other hand, highly urbanized cities like Singapore have successfully contained the virus (*COVID-19 may hit rural residents hard, and that spells trouble because of lack of rural health care*, 2020). Our data revealed a global negative association of percentage of rural population with COVID 19 mortality backed with the low death rate in Asia and Africa having high rural population percentage although no statistical significance was recorded. However, the finding was complimented with a significant high death rate recorded in North America (r=-0.665; p=0.005) comprising of low rural population. Similarly, Europe recorded a high death rate but not statistically significant with its low rural population. Contrasting the trend are Southeast Asian countries like Singapore and Japan and the Middle East countries with low rural population percentage and low mortality.

### Testing

The process of testing is directly associated with identification of more cases which in turn decreases the CFR (by increasing the denominator) and controls the viral spread by case isolation and other precautionary measures. Liang et al. demonstrated in their study that one additional test per 100 people led to 8% reduction in mortality rate, after controlling for case number, critical case rate, and various country-related factors (Liang *et al*., 2020).

We demonstrated a negative association of increased testing per million at the global scale. When calculated across various regions and continents, the Middle East countries (UAE, Bahrain, Israel, Qatar, Kuwait and Saudi Arabia) also showed association of more testing with less mortality; but no statistical relevance was reflected. While, Iran and Iraq from the same region had low testing associated with high mortality. South American countries on the contrary had high mortality rate which is attributed to their less testing practice (r=0.789; p=0,035). The deviation from the usual pattern was examined in the US with profound testing associated with low CFR but high mortality rate. Europe also tested more but showed high CFR as well as mortality. Asia and Africa tested less but reported low mortality comparatively. However, Singapore and South Korea from Asia had remarkably less mortality and high testing rate.

### GDP per Capita

An economically sound government is also essential for delivering quality public health services for the management, testing and containment of COVID-19 (Liang *et al*., 2020), (Stoller, 2020)as well as the swift execution of effective lockdown, quarantine and screening policies (Iacobucci, 2020),(Flaxman *et al*., 2020). R. Chaudhry et al., reported significant association of higher per capita GDP with poorer COVID 19 outcomes, increased critical cases and deaths per million population (Chaudhry *et al*., 2020). This may be explained by extensive testing, transparent and efficient surveillance and reporting systems in these countries or the increased accessibility to international travel, as travel has been documented as a major element contributing to viral spread (World Health Organization, 2020).

Our findings reflect a contrasting hypothesis, showing a global positive association of GDP with COVID mortality, which can be considered statistically significant with r=0.324 and p=0.051. While comparing the continent data, Europe and North America with high GDP reflected an increased mortality, however, only North America had a statistically significant association (r=0.481; p=0.059). The Indian subcontinent and Africa with low GDP had less mortality rate. Once again a reversed pattern was identified in Middle East countries and Southeast Asia (Singapore, Japan) with high GDP and less mortality. However, high mortality was reported in low GDP countries, like Iran and Iraq from the Middle East region.

### Health Expenditure

Percentage of GDP spent on healthcare by the government does not necessarily translate into developed healthcare system; it only reflects the attitude of the government towards the healthcare. It directly impacts the adequate supply of key medical equipment and supplies for the healthcare professionals as well as the COVID 19 patients (Peto, 2020; Ranney, Griffeth and Jha, 2020) (Adams and Walls, 2020). The novel character of COVID 19 highlights its specific diagnostic; containment as well as management requirements for which even the countries with advanced healthcare system were not prepared (Ranney, Griffeth and Jha, 2020). Hui Poh Goh et. al., have stated that the current health expenditure is not significantly associated with a high COVID-19 CFR depending upon the level of government preparedness in containing the pandemic(Goh *et al*., 2020). Such scientific studies are essential for mitigating risks and rational resource planning by policy makers leading to reduced the healthcare burden.

The global data of our study reveals a negative association of ‘health ependiture’ with COVID 19 mortality. However, the continent wise data once again negates the standard hypothesis and demonstrates a positive association, wherein, a low percentage of GDP spent on healthcare (<5%) in Middle East region was significantly associated with less mortality (r=0.616; p=0.043). Similar association was observed in the Indian subcontinent as well as Africa. On the other hand, high mortality was significantly associated with high health expenditure (>5%); like in North America (r=0.539; p=0.031). Similar findings were reported from Europe and South America. Excepting Japan, Afghanistan, Azerbaijan, Ukraine and Israel where more expenditure on healthcare by the government translated into less mortality.

### Beds

More lives can be saved if sufficient hospital beds are available, hence, number of hospital beds per 1000 people may serve as a proxy of access to public healthcare. Studies have confirmed that increasing the hospital bed capacity is vital to manage the healthcare demand in any future pandemic or emergency (Bedford *et al*., 2020; Liang *et al*., 2020) (Remuzzi and Remuzzi, 2020) (Hick and Biddinger, 2020).

We were not able to demonstrate a negative association of the ‘number of hospital beds per 1000 people’ in the global analysis. However, in the European continent, the high ‘number of hospital beds per 1000 people’ had an overall statistically significant and negative association with COVID mortality (r= −0.503; p=0.056). Low mortality rate was recorded in European countries with high number of such as Germany (8.3) and Poland (6.5). Low number of beds amounted to high mortality in UK (2.8) and Italy (3.4). Similarly, North America [Mexico (1.6) and USA (2.9)] had significant high mortality (r= −0.503; p=0.056) owing to low number of beds. In South America, Argentina (4.9) reported less mortality, whereas, Brazil (2.3) and Bolivia (1.1) experienced high mortality. However, Asia (except Japan) and Africa displayed a comparatively low mortality with regard to their less bed capacity. A record low mortality rate was observed with very high number of hospital beds in Japan (13.4).

### Lockdown Policy

The ravaging outspread of COVID-19 pandemic forced different countries to formulate strict policies to contain the virus. The time of implementation of the lockdown and preventive measures has remarkably been associated with the COVID outcomes irrespective of the economic status of a country (Anderson *et al*., 2020)-(*The early days of a global pandemic: A timeline of COVID-19 spread and government interventions*, 20210). China was able to suppress the virus spread in Wuhan, the epicenter of the pandemic, because of a stringent lockdown with early detection and isolation. Japan and South Korea on the other hand followed strict preventive policies (Banik *et al*., 2020). R. Chaudhry et al., stated that more time taken to seal the borders was positively associated the number of positive cases, and more stringent preventive measures was also positively associated with the number of recovered cases per million but did not limit the number of critical cases or mortality (Chaudhry *et al*., 2020).

### Diabetes mellitus

Attributes like pro-inflammatory and pro-coagulative state associated with diabetes play a critical role for worse COVID 19 prognosis(Apicella *et al*., 2017). The COVID 19 infection can aggravate severe metabolic complications through direct negative effects on β-cell function which might also lead to diabetic ketoacidosis in patients with diabetes, hyperglycaemia, unknown history of diabetes and potentially new-onset diabetes (Apicella *et al*., 2017). Recent studies reveal higher risk of severe/critical COVID 19 outcomes and mortality associated with diabetes (Mantovani *et al*., 2020)-(Barron *et al*., 2020).

In the present study, the dictum however does not hold true, as the global results displayed statistically significant but negative association between percentage of diabetes prevalence and COVID mortality (r= −0.384; p=0.019). Similarly, in the continent data analysis, North America [except USA (11%) and Mexico (14%)] displayed significant but negative association (r= −0.533; p=0.034). Even in Europe, countries with high diabetes prevalence like Portugal (10%) and Germany (10%) had less mortality, whereas, UK (4%) and Italy (5%) with low diabetes prevalence had high mortality. Similarly, despite high prevalence of diabetes in Asia comparatively fewer deaths were recorded (except Japan and Singapore which followed the positive association between diabetes and COVID 19 mortality). A positive association was revealed between diabetes and COVID 19 mortality in the Middle East region which was found to be statistically significant (r=0.647, p=0.031). This positive association was followed by Brazil (10%) and Chile (9%), where more deaths correlated to more diabetes prevalence, while, Argentina (6%) witnessed less deaths. In Africa, low mortality was examined in Ghana and Nigeria (both 3%) and high mortality in South Africa (13%) which was positively ascribed to diabetes prevalence.

### Smoking

Scientific evidence has revealed that smokers are more at risk of acquiring infectious diseases especially an array of respiratory disorders as smoking weakens the immune system(Tonnesen *et al*., 2019). In a systematic review, it was observed that smokers were 1.4 times more at risk of severe COVID-19 outcomes and approximately 2.4 times more at risk of admission to ICU, mechanical ventilation or death as compared to non-smokers(Vardavas and Nikitara, 2020). In a detailed study on smoking variable by Williamson et.al., higher risk of COVID 19 infection was observed in both present and former smokers (in models that were controlled for age and gender). However the observations reversed in fully controlled model which amounts to potential protective effect of smoking(Williamson *et al*., 2020). R. Chaudhryet. al., surprisingly demonstrated less distribution of critical cases and deaths in countries with higher smoking prevalence (Chaudhry *et al*., 2020). These results need to be verified to draw conclusions.

We were not able to infer any obvious pattern or significance for the smoking variable globally. In Asia, less mortality was recorded in India (11.5%) with low smoking prevalence, while Indonesia (39%) even with high smoking prevalence recorded less mortality. However, in the Middle East area, overall statistically significant but negative association between smoking and mortality was observed (r= −0.723; p=0.016) [Saudi Arabia (15%) and UAE (29%)]. In Europe, France (32%) and Spain (29%) reported high mortality, whereas, Germany (30%) and Poland (28%) with almost the same smoking prevalence reported low mortality. In North and South American continent, USA and Argentina both at 21.8% show a huge difference in mortality rate; Chile at 37% shows a high mortality rate, while, Brazil and Mexico even at 10-15% show high mortality.

### BCG Vaccination

Research literature reveals that Bacillus Calmette-Guerin (BCG) vaccination has beneficial effects on immunity; it not only provides a long lasting protective effect against tuberculosis, but also against many other infections including COVID-19 (Aronson *et al*., 2004)-(Curtis *et al*., 2020).

Our investigation of the global BCG vaccination policy disclosed that countries like the USA, Canada and Italy which did not have any universal BCG vaccination policy reported high COVID mortality. While, Poland and Portugal with universal BCG vaccination program reported less mortality. Asia and Africa where universal BCG program is running currently had a low mortality, however, in South America despite BCG program the mortality was high. Similar results were documented by Baniket al.(Banik *et al*., 2020), however, the protective potential of BCG vaccination was later put in doubts as the Asian and South American COVID 19 mortality started to shoot. World Health Organization on the other hand out rightly refutes the protective link of Bacille Calmette-Guerin (BCG) vaccine against COVID-19 (de Bree *et al*., 2018).

### Malaria

Some researchers have claimed that countries in Asian and African with high prevalence of malaria have relatively fewer coronavirus cases and mortality. The WHO has strongly responded against such hypothesis stating that these findings were only based on the total number of COVID-19 cases as of March 17, 2020.

We observed that in Asia and Africa where malarial prevalence is quiet high, the COVID 19 mortality was low. Being a ‘no malarial zone’ Europe reported high mortality. However, in North and South American continent (except USA and Canada) malarial prevalence as well as mortality both were high.

### Communicable Diseases

It has been proposed that the people of countries with high prevalence of communicable disease such as, malaria, tuberculosis and HIV might be immunologically more protected against infectious diseases(Hogan *et al*., 2020). This hypothesis, once again has been out rightly overruled by WHO. To prevent any harm due to the misconception regarding the protective association of communicable diseases, WHO highly recommends the maintenance of prevention activities and health-care services for all common chronic communicable diseases, to reduce the morbidity and mortality of COVID-19.

We observed that percentage of deaths due to communicable diseases and poor nutrition conditions, in the population were surprisingly negatively associated with the COVID mortality, when assessed all over the world. Accordingly, Africa which has a huge communicable disease burden was strikingly spared of COVID 19, when compared to developed regions like Europe USA and Canada which are badly hit by COVID 19 and have low communicable disease prevalence. Similarly, low mortality in Asia was significantly attributed to its high communicable disease prevalence (r= −0.436; p=0.043). Middle East countries are the deviations in the trend which have low communicable disease prevalence and yet low mortality; and South America with a reverse of high mortality and high communicable disease prevalence.

### Limitations

There are a few obvious limitations of the study that need to be acknowledged. The study is being conducted in the midst of the pandemic and as such the data is subject to change over any plausible short duration. However, this could serve as the strength of study, wherein our results will reflect an overview of the global impact of the virus. Another important confounding factor that impacts the mortality rate and our study observation is how the COVID-19 mortality is calculated. Currently different countries have different methods and criteria for analyzing their respective mortality.

## Conclusion

Our findings indicate that there is no set of factors which follow a specific pattern across the globe. Our observations once again revealed a vast variation in the COVID mortality across the globe. However, we observed a continent and region wise significant association of some factors with the COVID mortality. The interaction between the factors analyzed will provide an insight into the confounding effects and the complexity of the disease. The observations of our study will serve as a evidence based data for generating predictive model for the severe COVID 19 outcomes and death. We hope the extensive data analysis will help in risk stratification of most vulnerable population for revising the COVID 19 containment and management action plan. Much still needs to be learned about the novel COVID-19 which has impacted all domains of human civilization. COVID 19 has not only challenged the intellect of human race but also their ability to fight the unseen enemy.

## Data Availability

Nil

## Contribution details

**Contribution details:**
| S.no | Concept, | Design, | Definition of intellectual content, | Literature search, | Data acquisition | Data analysis, | Statistical analysis, | Manuscript preparation, | Manuscript editing | Manuscript review. |
| --- | --- | --- | --- | --- | --- | --- | --- | --- | --- | --- |
| 1 | *Vineet Jain* | Yes | Yes | - | - | Yes |  | Yes | Yes | Yes |
| 2 | *Nusrat Nabi,* |  | Yes | Yes | Yes |  |  | Yes | Yes | Yes |
| 3 | *Kailash Chandra,* | Yes | Yes | - | Yes | Yes | Yes |  | Yes | Yes |
| 4 | *Sana Irshad,* | - | - | Yes | Yes | Yes |  | Yes | Yes | - |
| 5 | *Varun kashyap* | - | Yes | Yes |  | Yes | Yes |  | Yes | Yes |
| 6 | *Sunil Kohli,* | Yes | Yes | - | - | - | - | - | Yes | Yes |
| 7 | *Arun Gupta* | Yes | Yes | - | - | Yes | - | - | Yes | Yes |

